# DETERMINANTS OF BIRTH PREPAREDNESS AND EMERGENCY READINESS AMONG PREGNANT WOMEN ATTENDING ANTENATAL CLINIC IN SECONDARY AND TERTIARY HOSPITALS IN PORT HARCOURT METROPOLIS

**DOI:** 10.1101/2025.09.03.25334954

**Authors:** Kingsley Uwaezuoke Obi, Emmanuel Etim Clement, Best Ordinioha

## Abstract

This study aimed to identify determinant of Birth Preparedness and Emergency Readiness (BPER) among pregnant women in the Port Harcourt Metropolis of Rivers State, Nigeria. The study adopted a comparative cross-sectional design using a multi-staged clustered sampling technique to select 1,084 participants from secondary and tertiary health facilities in Port Harcourt. Semi-structured interviewer-administered questionnaire was used to gather data on the determinants and socio-demographic factors. Statistical analysis was conducted to examine associations between socio-demographic factors and BPER and practice. The study revealed that the average age of the participants in the secondary health facility was predominantly 25-29 years (54.45%) compared to 45.2% in the tertiary health facility, with a statistically significant difference (p = 0.0001). Most respondents in both health facilities had secondary education, with 63.3% in the secondary facility and 69.0% in the tertiary facility, showing a significant difference (p = 0.0001). A greater proportion of participants in the secondary facility were unemployed (35.6%) compared to the tertiary facility (42.4%), also with a statistically significant difference (p = 0.0001). A large majority of respondents in both groups were Christian, with 98.3% in the secondary health facility and 97.8% in the tertiary health facility, showing no significant difference (p = 0.509). Findings further revealed that participants who had not experienced a miscarriage or pregnancy loss were statistically significantly associated with better practice of BPER in the bivariate model. Also, 84.9% of respondents in the secondary health facility had poor practice, compared to 77.1% in the tertiary health facility, with a statistically significant difference (p = 0.001). Key covariates, such as a history of miscarriage, complications during pregnancy, and multiple pregnancies, showed statistically significant associations with the practice of BPER (p = 0.0001).

## Introduction

Globally, an estimated 800 women die each day from complications related to pregnancy and childbirth. The burden of these maternal deaths is disproportionately high in developing nations, which account for 99% of all cases, with over half occurring in the sub-Saharan African region (World Health Organization, 2016). A primary intervention for reducing maternal mortality is birth preparedness, a comprehensive strategy designed to ensure access to skilled care at delivery.

The Birth Preparedness and Complication Readiness (BP/CR) strategy is a key component of this approach. It empowers women and their families to plan for a safe birth and anticipate responses to obstetric emergencies. Key elements include: recognizing danger signs; identifying a preferred birth location and skilled birth attendant; arranging transportation to an emergency care facility; securing funds for associated costs; designating a companion for labor and delivery; and identifying potential blood donors (World Health Organization, 2010). This planning is intended to facilitate rapid decision-making and mitigate the critical delays in seeking care that contribute to maternal mortality.

International efforts, particularly during the Millennium Development Goal (MDG) era, yielded significant progress, achieving a 44% reduction in the global maternal mortality ratio (MMR). The subsequent Sustainable Development Goals (SDGs) aim to build on this by further reducing the global MMR by more than two-thirds by 2030, effectively aiming to end preventable maternal deaths (UNICEF, 2010; WHO, 2014). The BP/CR approach, developed within the framework of the Safe Motherhood Initiative and endorsed by the World Health Organization as part of its Focused Antenatal Care (FANC) model, is recognized as a critical strategy for achieving this target (JHPIEGO, 2004; Karkee, Lee & Binns, 2014; Ijang et al., 2019).

The practice of BP/CR is the behavioral manifestation of knowledge and attitudes. It encompasses the specific actions women take, such as making delivery arrangements and identifying healthcare providers, which are directly influenced by their understanding of risks and their attitude towards preparedness (Mason et al., 2012). These practices are, in turn, shaped by a range of sociodemographic and determinant factors. Key sociodemographic variables include age, educational attainment, marital status, and socioeconomic status. Determinants encompass broader external and internal factors such as healthcare access, cultural beliefs, previous birth experiences, and level of family support (Hussein & Hadi, 2010). These factors collectively influence a woman’s capacity to translate knowledge into effective practice, ultimately impacting maternal and neonatal health outcomes.

Despite its established importance, research indicates that low awareness of obstetric danger signs and poor BP/CR practice remain significant barriers, particularly in rural areas, leading to delays in seeking care (Ekabua et al., 2011; Nsemo & Offiong, 2016). Nationwide surveys attribute low levels of skilled birth attendance and preparedness to systemic, socioeconomic, and cultural obstacles, including financial constraints, distance to health facilities, the perception that facility-based delivery is unnecessary, and the sudden onset of labor (National Population Commission [NPC], 2019; 2014). It is against this backdrop that this study intends to identify and compare the determinants and socio-demographic factors influencing the practice of Birth Preparedness and Emergency Readiness (BPER) among pregnant women in the Port Harcourt Metropolis of Rivers State, Nigeria.

## Aim and Objectives of the Study

This study aimed to identify determinants of Birth Preparedness and Emergency Readiness (BPER) among pregnant women in the Port Harcourt Metropolis of Rivers State, Nigeria. The following specific objectives guided the study:

1. To identify determinants that affect the practice of BPER among antenatal clinic attendees in secondary and tertiary healthcare facilities.
2. To identify the relationship between socio-demographic characteristics, Obstetrics and Gynaecology history of the antenatal clinic attendees in secondary and tertiary healthcare facilities and their practice of BPER.

## Research Questions

The following understated research questions guided this study:

1. What are the determinants that affect the practice of BPER among antenatal clinic attendees in secondary and tertiary healthcare facilities?
2. What is the relationship between socio-demographic characteristics, Obstetrics and Gynaecology history of the antenatal clinic attendees in secondary and tertiary healthcare facilities, and their practice of BPER?

## Methods and Materials

This study employed a comparative cross-sectional design and was conducted in Rivers State, Nigeria, to identify the determinants of Birth Preparedness and Emergency Readiness (BPER) among pregnant women in the Port Harcourt Metropolis. The population of the study comprised pregnant women attending antenatal clinics of the selected secondary and tertiary healthcare facilities in Port Harcourt metropolis. The pregnant women in Port Harcourt Metropolis, Rivers State, represent a diverse and dynamic group that plays a key role in the overall maternal health landscape of Nigeria. Consenting pregnant women and women who have attended at least three antenatal clinic visits with a gestational age of ≥12 weeks were included, while first-time pregnant women who are attending ANC visits for the first time were excluded because they have no previous experience. Also excluded were pregnant women who were critically ill during the period of the study. The sample size of this study was calculated using a comparative cross-sectional study design for two proportions by Cochrane;

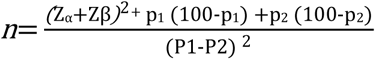

Where;

n= minimum sample size required for each group

<u>Z_α_</u> = standard normal deviation (at confidence level of 95% =1.96)

<u>Z_β_</u> = standard normal deviation for statistical power (at 80% = 0.84)

P1 = 46.8% (proportion of pregnant women who had good practices of BPER in a tertiary facility by Deji et al., 2021).

P2 = 44.9%% (proportion of pregnant women with complication readiness in secondary facility by Anikwe et al., 2020).

P1-P2= difference between the two proportions.

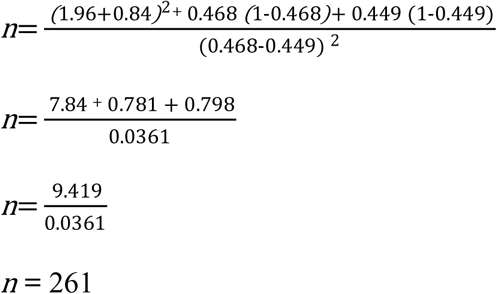

Considering 10% non-response rate,

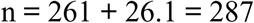

Applying the design effect of 2, approximately, the minimum sample size for this study

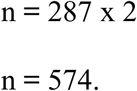

A total of 1,148 respondents were recruited for the study, with 574 participants each from selected secondary and tertiary healthcare facilities in Port Harcourt Metropolis. A multi-stage clustered sampling technique was used: the metropolis was first stratified into two LGAs, from which four secondary facilities (two per LGA) were randomly selected, while the two public tertiary facilities were purposefully chosen. Systematic sampling was employed to select respondents, using a calculated sampling interval (K = N/n), where N is average daily clinic attendance and n is the sample size. The first participant was randomly selected, followed by every nth consenting attendee. Data were collected using a semi-structured, interviewer-administered questionnaire adapted from previous studies and tailored to the study’s scope. Section one captured sociodemographic and reproductive history, while section two assessed which explored the determinants of BPER. The instrument was peer-reviewed by Public Health experts and pre-tested on 5% of the sample at a primary healthcare facility in Ikwerre LGA to ensure reliability. Data collection spanned six months. Trained research assistants, selected for their communication skills and familiarity with the study area, administered the questionnaires during designated ANC clinic days. Completed forms were immediately reviewed for completeness. Data were coded and entered into Microsoft Excel, then transferred to SPSS version 25 for analysis. Chi-square tests were applied to examine associations between BPER status (prepared vs. unprepared) and socio-demographic and reproductive health variables. Statistical significance was set at p < 0.05.

## Results

The table reveals that 25-29yrs is the average age of more than half 295(54.45%) of the correspondents in the secondary health facility compared to a lesser number 245(45.2%) of the participants in the tertiary health facility with a statistically significant difference at *p*=(0.0001). Majority 343(63.3%) of the respondents in the secondary health facility compared to more 374(69.0%) of the tertiary health facility participants has attained secondary education with a statistically significant difference at p=(0.0001). There was no statistically significant difference in the marital status among respondents in the secondary and tertiary health facility are married. Many 193(35.6%) secondary health facility participants compared to more 230(42.4%) of the tertiary health facility participants are unemployed with a statistically significant difference at p=(0.0001). Christianity is the religion of most 533(98.3%) of the respondents in the secondary health facility compared to a lesser fraction 530(97.8%) of the participants in the tertiary health facility with no statistically significant difference at p=(0.509). Among the Christians, many 166(30.6%) of the respondents in the secondary health facility are protestants compared to more than half 306(56.5%) of the tertiary health facility respondents with statistically significant difference at p=(0.0001). More than half 286(52.8%) of the respondents in the secondary health facility compared to a lesser fraction 19(35.8) of the participants in the tertiary health facility are of 3 gravidities with a statistically significant difference at p=(0.0001).

The table shows that majority 346(63.8%) of the respondents in the secondary health facility compared to a lesser number 217(40.0%) of the tertiary health facility participants has 1 parity with a statistically significant difference at p=(0.0001). More than half 288(53.1%) secondary health facility participants compared to many 264(48.7%) of the tertiary health facility participants are in there second trimester with a statistically significant difference at p=(0.0001).

Table 2a shows that majority 268(49.4%) of the respondents in secondary health facility compared to more 386(71.2%) of the respondents in tertiary health facility have not experienced a miscarriage or pregnancy loss with a statistically significant difference at *p*=(0.0001). A larger number 367(67.7%) of secondary health facility respondents compared to less number 253(46.7%) tertiary health facility respondents have had complications in previous pregnancy with a statistically significant difference at *p*=(0.0001). No complication was experienced by fewer 175(32.3%) respondents in secondary health facility compared to more than half 289(53.3%) of the respondents in tertiary health facility with a statistically significant difference at *p*=(0.0001).

**Table 1a:**
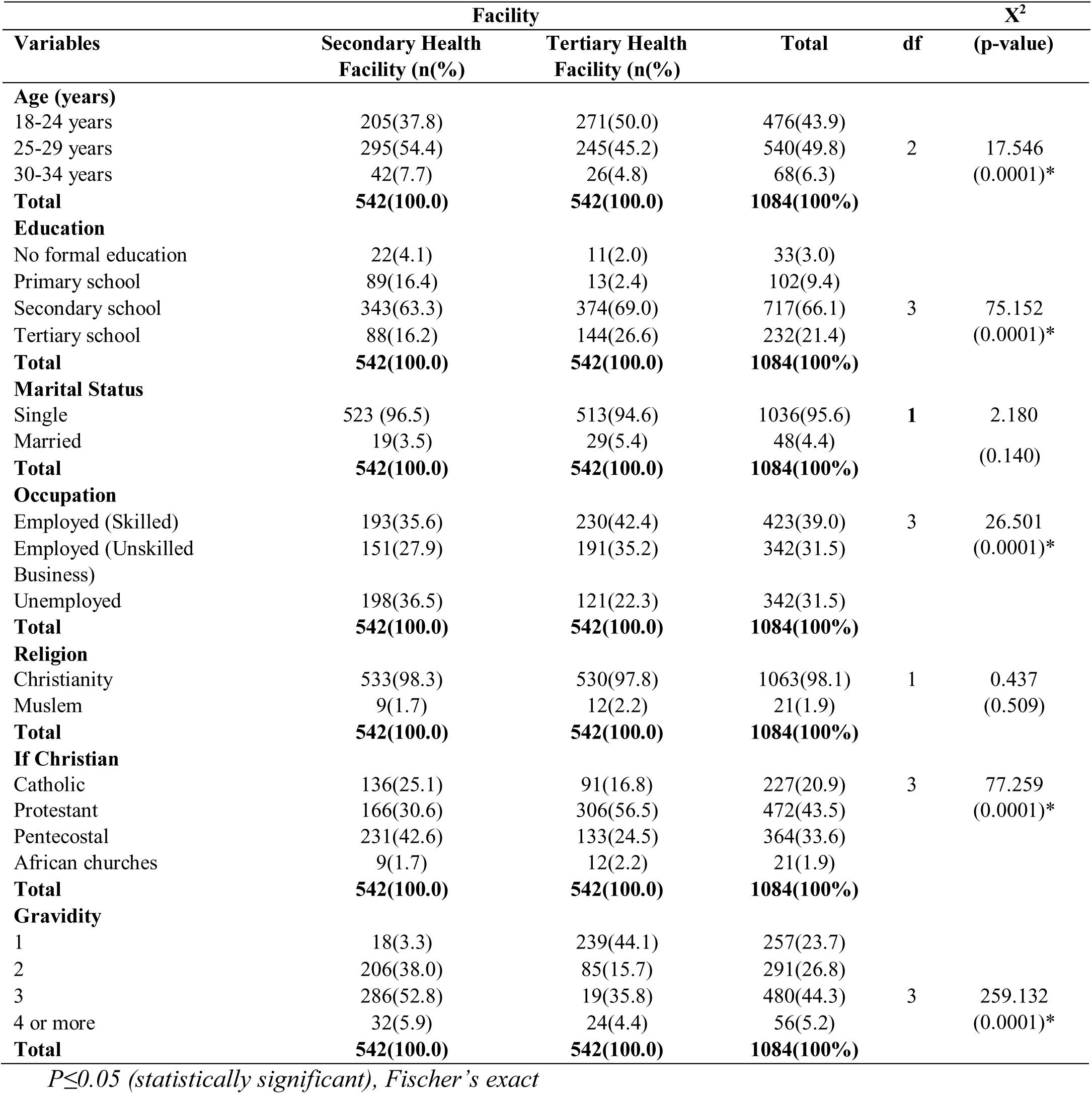
Socio-Demographic Characteristics of Respondents

**Table 1b:** Socio-Demographic Characteristics of Respondents

| Variables | Facility |  | Total | df | (p-value) |
| --- | --- | --- | --- | --- | --- |
|  | Secondary Health Facility (n(%)) | Tertiary Health Facility (n(%)) |  |  |  |
| **Parity** |  |  |  |  |  |
| None | 9(1.7) | 215(39.7) | 224(20.7) | 2 | 238.967<br>(0.0001)* |
| 1 | 346(63.8) | 217(40.0) | 563(51.9) |  |  |
| 2 | 187(34.5) | 110(20.3) | 297(27.4) |  |  |
| **Total** | **542(100.0)** | **542(100.0)** | **1084(100%)** |  |  |
| **Gestational age** |  |  |  |  |  |
| 1st trimester | 91(16.8) | 156(28.8) | 247(22.8) | 2 | 24.047<br>(0.0001)* |
| 2nd trimester | 288(53.1) | 264(48.7) | 552(50.9) |  |  |
| 3rd trimester | 163(30.1) | 122(22.5) | 285(26.3) |  |  |
| **Total** | **542(100.0)** | **542(100.0)** | **1084(100%)** |  |  |

**Table 2a:**
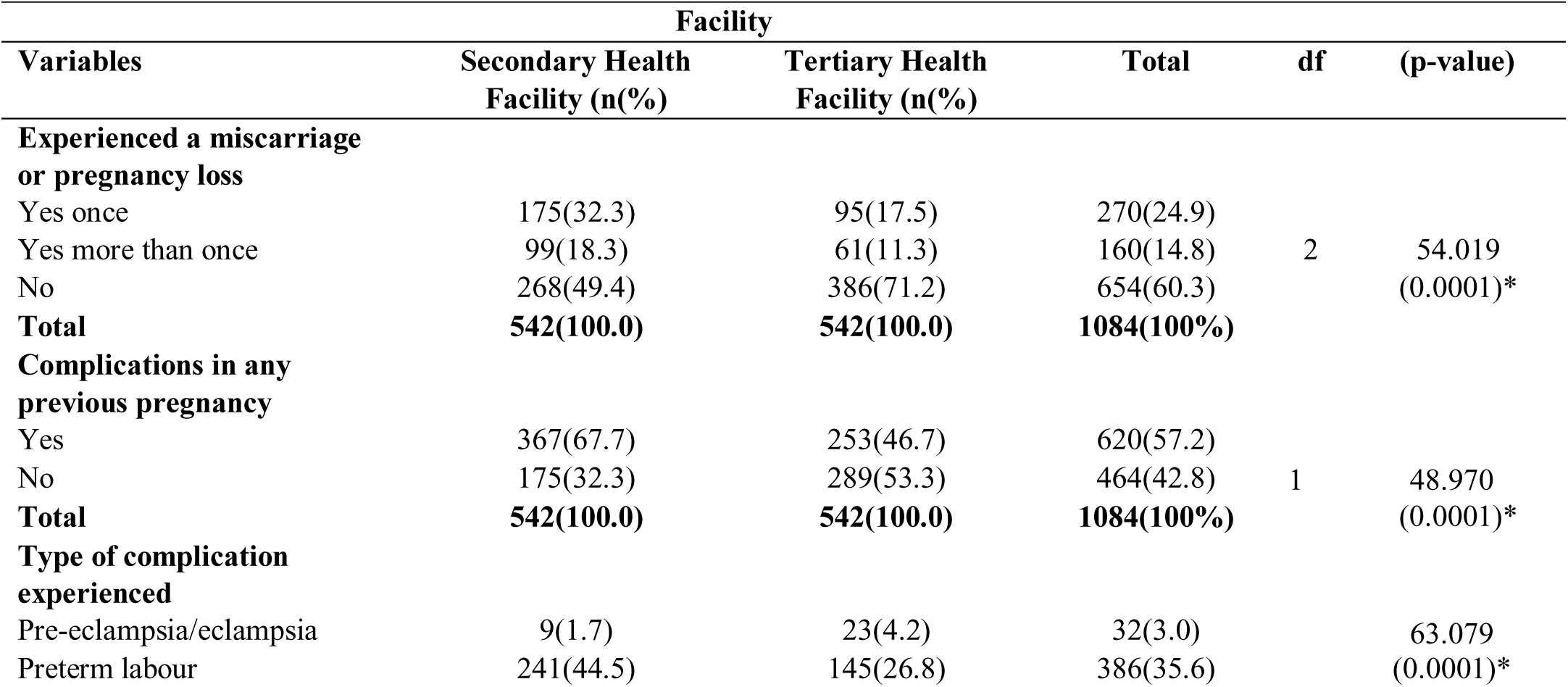
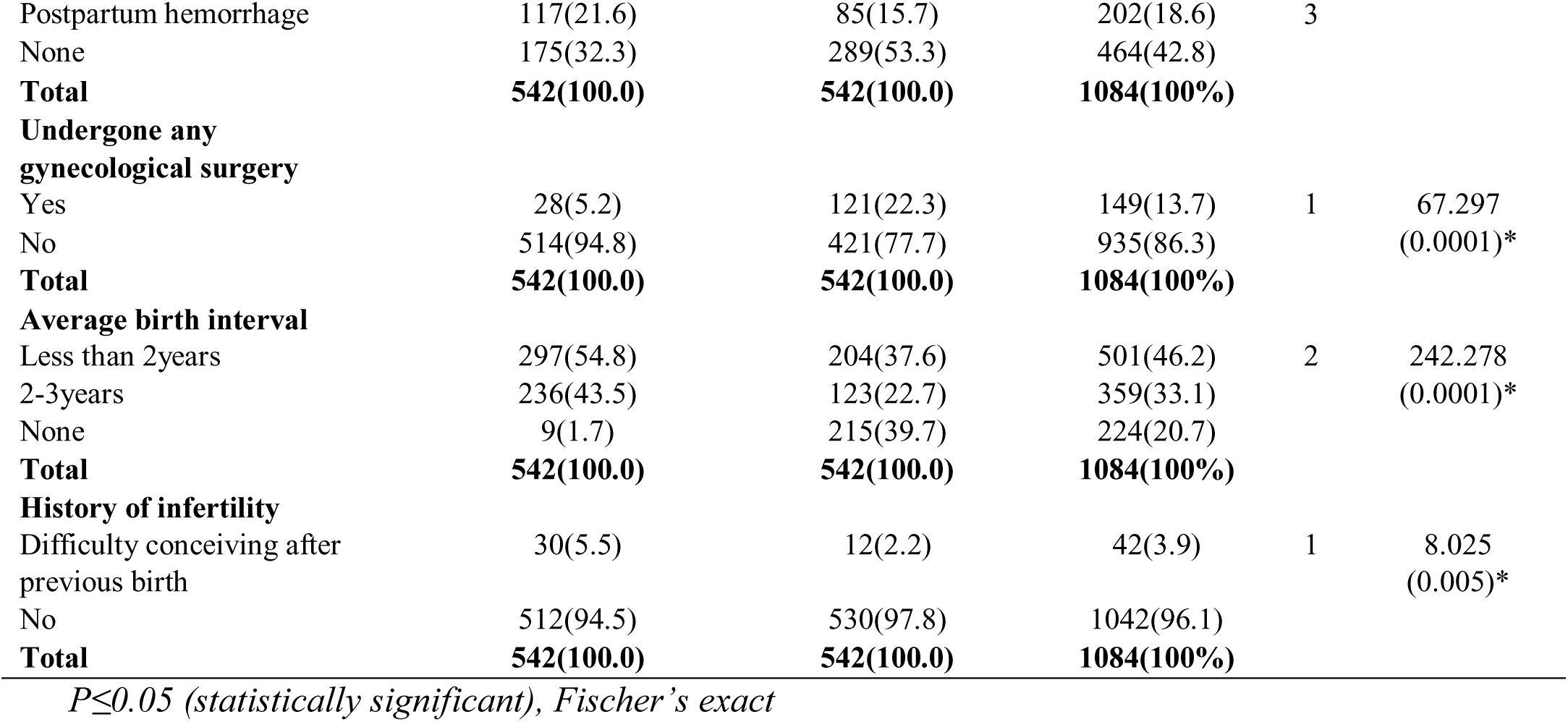
History of Obstetrics/Gynaecology

Most 514(94.8%) of the respondents in the secondary health facility compared to majority 421(77.7%) of the tertiary health facility participants have undergone gynecological surgery with a statistically significant difference at p=(0.0001). Less than 2years was the average birth interval for more than 297(54.8%) secondary health facility participants compared to fewer number 297(54.8%) of the tertiary health facility participants with a statistically significant difference at p=(0.0001). There was no history of infertility for most 512(94.5%) of the participants in secondary health facility compared to a higher fraction 530(97.8%) of the respondents in the tertiary health facility.

Table 2b indicates that most 457(84.3%) of the respondents in secondary health facility compared to more than half 50(10.8%) of the respondents in tertiary health facility were not diagnosed with any medical conditions with a statistically significant difference at *p*=(0.0001). A larger fraction 533(98.3%) of secondary health facility respondents compared to most 530(97.8%) tertiary health facility respondents do not have a history of multiple pregnancy with no statistically significant difference at *p*=(0.509). Vaginal delivery was the mode of delivery for many 192(35.4%) of the respondents in secondary health facility compared to a lesser number 171(31.5%) of the respondents in tertiary health facility with a statistically significant difference at *p*=(0.0001). There is statistically significant difference between participants in the secondary and tertiary health facility who attend or do not attend antenatal clinic regularly for every scheduled appointment. at p=(0.048).

**Table 2b:**
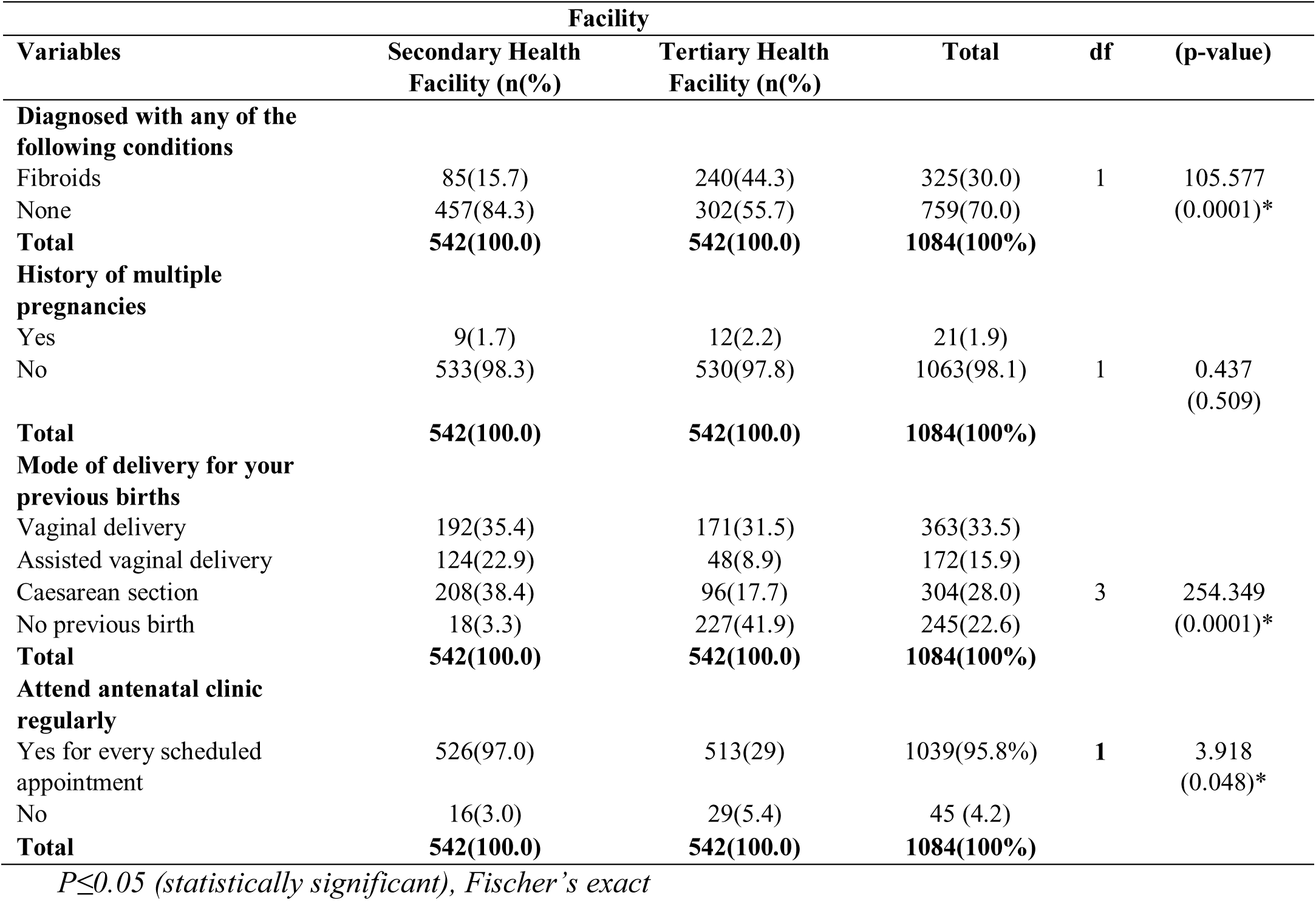
History of Obstetrics/Gynaecology

Table 3 shows that covariates; those who did not experienced a miscarriage or pregnancy loss by participants was statistically significantly associated with practice of BPER among ANC attendee in the bivariate model at (p=0.0001) (OR=4.606, 95% CI: 2.442-7.691). The result showed that there was a statistically significant association between having complications and with practice of BPER among ANC attendee in the bivariate model with (p=0.014) (OR=3.661, 95% CI: 0.014-1.989). Undergone any gynecological surgery had no statistically significant association with practice of BPER among ANC attendee in the bivariate model at (p=0.995). Similarly, history of infertility had no statistically significantly association with practice of BPER among ANC attendee in the bivariate model at (p=0.997). The result showed that history of multiple pregnancies had a statistically significant association with ANC attendee in the bivariate model at (p=0.0001) (OR=30.110, 95% CI: 0.044-0.277). Also, mode of delivery for your previous births had statistically significant association with ANC attendee in the bivariate model at (p=0.0001).

**Table 3:** Determinants that affect the practice of BPER among ANC attendees

| Parameter | Response Category | Odds Ratio | p-value | 95% C.I. of Odds Ratios |  |
| --- | --- | --- | --- | --- | --- |
|  |  |  |  | Lower | Upper |
| **Experienced a miscarriage or pregnancy loss** | Yes once | ref |  |  |  |
|  | Yes more than once | 3160711649.057 | 0.993 | 0.000 | - |
|  | No | 4.606 | 0.0001* | 2.442 | 7.691 |
| **Had complications** | Yes | ref |  | 0.014 | 1.989 |
|  | No | 3.661 | 0.014 * | - | - |
| **Undergone any gynecological surgery** | Yes | ref |  | 0.000 | - |
|  | No | 456499063.961 | 0.995 | - | - |
| **History of infertility** | Difficulty conceiving after previous birth | ref |  | 0.000 | - |
|  | No | 398071552.186 | 0.997 | - | - |
| **History of multiple pregnancies** | Yes | ref |  | 0.044 | 0.277 |
|  | No | 0.110 | 0.0001 | - | - |
| **Mode of delivery for your previous births** | Vaginal delivery | ref |  |  |  |
|  | Assisted vaginal delivery | 190055863.865 |  | 0.000 | - |
|  | Caesarean section | 0.251 | **0.0001** | 0.165 | 0.382 |
|  | No previous birth |  | 0.994 |  |  |

*( $P \leq 0.05$ = statistically significant); (ref=reference)*

Table 4 shows that covariates; age (30-34 years) of the participants was statistically significantly associated with practice of BPER among ANC attendee in the bivariate model at (p=0.0001) (OR=4.606, 95% CI: 2.697-7.865). The result showed that there was a statistically significant association between level of education completed and with practice of BPER among ANC attendee in the bivariate model with (p=0.0001) (OR=3.661, 95% CI: 2.177-6.157). The occupation had a statistically significant association with the practice of BPER among ANC attendees in the bivariate model at (p=0.0001) (OR=204.130, 95% CI: 28.291-1472.8). Similarly, religion no statistically significantly association with practice of BPER among ANC attendees in the bivariate model at (p=0.058). The result showed that parity had no statistically significant association with ANC attendance in the bivariate model at (p=0.631). Also, gestational age had no statistically significant association with ANC attendance in the bivariate model at (p=0.0001).

**Table 4:**
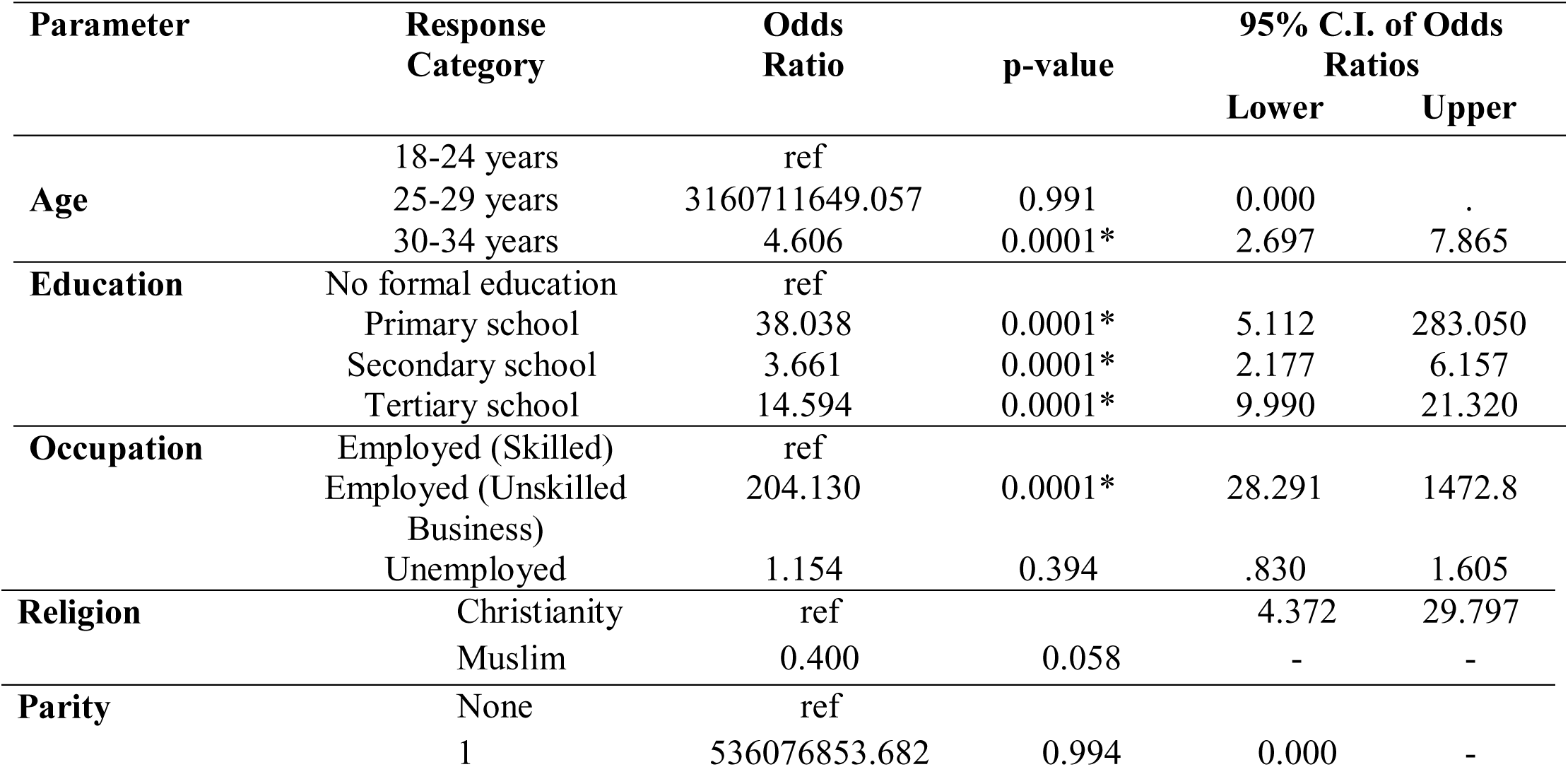
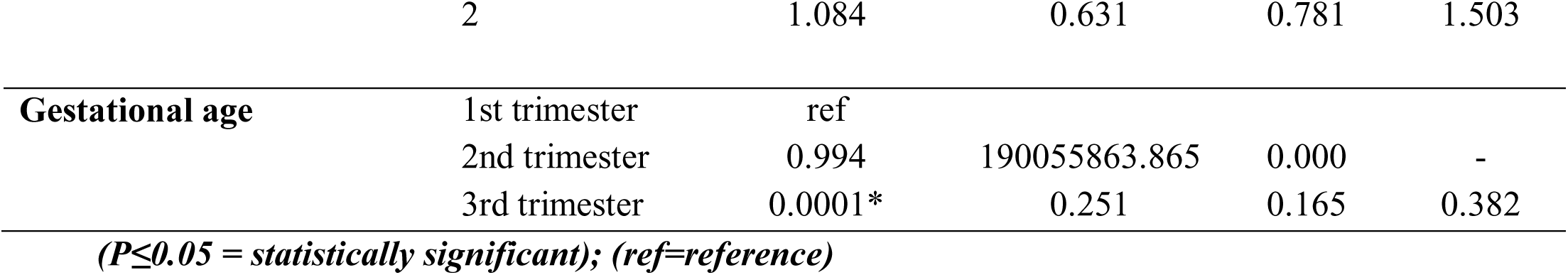
Socio-demographic characteristics of BPER among ANC attendee

## Discussion of Findings

### Determinants that affect the practice of BPER among antenatal clinic attendees in secondary and tertiary healthcare facilities

Findings showed that participants who had not experienced a miscarriage or pregnancy loss were statistically significantly associated with better practice of BPER in the bivariate model. This finding is consistent with the study by Aduloju et al. (2017) conducted in a Teaching Hospital in South West Nigeria, which reported that higher parity (p < 0.05) was a significant determinant of birth preparedness and emergency readiness practices. However, it contrasts with the findings of Opu et al. (2025) in Eastern Uganda, where previous miscarriage experience was not found to influence birth preparedness and emergency readiness practices among pregnant women attending antenatal clinics. This specific variable was not directly addressed in the studies by Mahato (2019), Mahore (2023), or Sabo et al. (2024). However, Mahato reported that multiple ANC visits and family involvement in decision-making significantly increased facility-based deliveries, which may indirectly relate to better BPER practices among women with less traumatic obstetric histories. While direct comparison is limited, both studies support the view that positive reproductive history and family support encourage better maternal care practices. A statistically significant association was observed between having experienced complications and BPER practice. In line with this, Sabo et al. (2024) reported that knowledge of pregnancy danger signs was significantly associated with ANC service utilization. This implies that women with complication experience or awareness are more likely to be proactive about maternal care, similar to findings in the current study. Consistent with previous findings past complications increase awareness and preparedness for future births. No statistically significant association was found between gynecological surgery history and BPER practice. This factor was not considered in Mahato (2019), Mahore (2023), or Sabo et al. (2024), making direct comparison difficult. However, gynecological surgical history may not be a widely recognized influencer of birth preparedness behaviors unless directly linked with pregnancy outcomes. There is no clear agreement, but both current and previous studies suggest other factors (e.g., complications, education, support) may be more influential than surgical history.

### Relationship between Socio-demographic characteristics and practice of BPER among ANC attendees

The current findings from this study demonstrate that certain sociodemographic variables such as age (30–34 years), education level, and occupation are statistically significantly associated with the practice of Birth Preparedness and Emergency Readiness (BPER) among antenatal care (ANC) attendees, while other factors like religion, parity, and gestational age were not significantly associated. Age (specifically 30–34 years) was significantly associated with the practice of BPER. Aduloju et al. (2017) similarly found that older age was significantly associated with better birth preparedness and complication readiness (BPER). Mandah & Peter-Kio (2025) also supported that obstetric correlates, often influenced by age, were associated with BPER. There is consistency between both current and previous studies support that age (especially older maternal age) is positively associated with better BPER practices. There was a statistically significant association between educational level completed and BPER practice, Rustagi et al. (2021) showed higher knowledge of ANC among more educated women. Bejitual and Jabessa (2021) found that secondary or tertiary education was strongly associated with BPER practice (e.g., OR=4.12 for college graduates). Aduloju et al. (2017) noted that tertiary education of women significantly predicted better preparedness. Anikwe et al. (2020) also supported that early booking, often linked to educational awareness, predicted better knowledge. There is an agreement that education level is a strong predictor of BPER across studies. Occupation was found to be significantly associated with BPER practice. Bejitual & Jabessa (2021) reported maternal occupation as a significant independent predictor of BPER Aduloju et al. (2017) found that both the woman’s and her spouse’s employment were associated with better BPER outcomes. There is consistency that occupational status contributes to economic empowerment and awareness, which promotes BPER practice.

Religion had no statistically significant association with BPER practice, this is not in line with findings by (Rustagi, Aduloju, Bejitual) who reported religion as a significant predictor of BPER. However, most focused on education, occupation, parity, and ANC service use. This indicates that religion to play a limited role in influencing BPER practices across multiple. The parity of the respondents was not statistically significantly associated with BPER practice. This sis similar to reports of Aduloju et al. (2017) that higher parity was significantly associated with BPER, possibly due to accumulated pregnancy experiences. Also, Bejitual & Jabessa (2021) mentioned that having 1–3 prenatal visits or more was significantly associated with BPER, though parity itself was not emphasized. The finding by Mandah & Peter-Kio (2025) indirectly suggest a relationship between obstetric history and preparedness. Current result differs from Aduloju et al. who found higher parity positively associated with preparedness. This inconsistency may be due to contextual or sample-specific factors such as health education access or cultural norms. Gestational age showed no statistically significant association with BPER. Finding by Anikwe et al. (2020) emphasized early booking (1st or 2nd trimester) as a significant predictor of preparedness and knowledge, implying that gestational age at ANC initiation could matter. Other studies such as Bejitual and Jabessa (2021) linked frequency of prenatal visits (which increases with gestational age) to BPER practice. While gestational age itself may not always show a direct correlation, timing of ANC registration and visit frequency which relate to gestational age are often indirectly significant in previous studies.

## Conclusion

In conclusion, sociodemographic factors such as age, level of education, occupation, and previous obstetric experiences (history of complications, multiple pregnancies, or previous mode of delivery) were found to be significantly associated with BPER practices. In contrast, variables such as religion, parity, gestational age, and history of infertility showed no significant influence. Overall, this study underscores the urgent need for targeted educational interventions, improved communication between healthcare providers and ANC clients, and context-specific strategies to bridge the preparedness gap, particularly in secondary health facilities. Addressing socioeconomic barriers and enhancing ANC service quality are critical to improving maternal outcomes through better birth preparedness and emergency response.

## Recommendations

Based on the findings and conclusion of this study, the following recommendations were proffered:

1. Targeted interventions should be developed for subgroups statistically associated with poor BPER practices, such as women with lower educational attainment, younger mothers, and those with no prior birth complications.
2. Regular assessments should be implemented to track progress on BPER indicators at both secondary and tertiary health facilities, ensuring evidence-based adjustments to programs and policies.

## Data Availability

All data used are available in the manuscript

